# Reproducibility of Apparent Diffusion Coefficient and Restriction Spectrum Imaging Restriction Score in the Prostate Across MRI Sessions, Vendors, and Acquisition Settings: a Prospective Study

**DOI:** 10.64898/2026.05.10.26352843

**Authors:** Yuze Song, Christopher C Conlin, Kang-Lung Lee, Anna M Dornisch, Tristan Barrett, Son Do, Deondre D Do, Daniel JA Margolis, Rebecca Rakow-Penner, Anders M Dale, Michael A Liss, Tyler M Seibert

## Abstract

**Background:** Diffusion-weighted MRI is central to prostate cancer detection, but apparent diffusion coefficient (ADC) has limited reproducibility across scanners and sites. Restriction Spectrum Imaging restriction score maximum value (RSIrs-max) may provide a more reproducible biomarker.

**Purpose:** To evaluate cross-session reproducibility of within-lesion mean ADC and RSIrs-max on prostate MRI, including same-vendor and cross-vendor comparisons, and in unfavorable-histology prostate cancer (uhPC) and different interpolation settings.

**Materials and Methods:** In this prospective study, participants with suspected or known prostate cancer enrolled from August 2022 to January 2026 underwent two MRI examinations including an RSI protocol. MRI-visible lesions were contoured on T2-weighted MRI; in participants with multiple lesions, the index lesion was selected. Mean ADC and RSIrs-max were measured within MRI-visible lesions. Analyses included all visible lesions, same-vendor and cross-vendor subgroups, participants with uhPC, and 20 participants with scans reconstructed with and without zero-filled interpolation (a setting with different defaults across vendors). Pearson correlation coefficients with 10,000 bootstrap resamples were used to estimate 95% confidence intervals.

**Results:** Sixty-one male participants (median age, 69 years [IQR, 63-74]) were evaluated; 58 of 61 (95%) had MRI-visible lesions, and 26 of 58 (45%) had uhPC. For all MRI-visible lesions, correlations were 0.55 (95% CI: 0.23-0.76) for mean ADC and 0.83 (95% CI: 0.72-0.90) for RSIrs-max. In same-vendor scans, correlations were 0.76 (95% CI: 0.27-0.95) and 0.88 (95% CI: 0.72-0.96); in cross-vendor scans, they were 0.31 (95% CI: −0.07-0.62) and 0.79 (95% CI: 0.65-0.89), respectively. In uhPC, correlations were 0.42 (95% CI: −0.02-0.83) for mean ADC and 0.90 (95% CI: 0.77-0.96) for RSIrs-max. With inconsistent versus consistent interpolation, RSIrs-max correlation increased from 0.73 (95% CI: 0.48-0.89) to 0.89 (95% CI: 0.78-0.96).

**Conclusion:** ADC showed limited reproducibility, particularly across vendors. RSIrs-max has stronger between-session reproducibility across same-vendor, cross-vendor, uhPC, and interpolation analyses.

## Introduction

Magnetic resonance imaging (MRI) plays a key role in the early diagnosis of prostate cancer. As recommended by the National Comprehensive Cancer Network (NCCN) and European Association of Urology (EAU) guidelines, the standard diagnostic pathway includes multiparametric MRI (mpMRI) prior to biopsy, which has been shown to reduce the number of unnecessary biopsies and improve the detection of clinically significant prostate cancer (csPCa) ^1–7^. In mpMRI, diffusion-weighted imaging (DWI) plays a central role in the detection of csPCa, particularly in the peripheral zone where the incidence of cancer is highest ^8,9^. Despite the success of DWI for diagnosing prostate cancer, there are concerns about limited reproducibility that can introduce variability into both diagnostic assessment and downstream clinical decision-making ^10^. Apparent diffusion coefficient (ADC) is a quantitative parameter derived from DWI that is widely used to evaluate suspected prostate cancer lesions. A recent meta-analysis suggested ADC within lesions on prostate MRI has independent prognostic value beyond clinical information in patients undergoing prostatectomy ^11^. However, ADC measurements vary substantially across different MRI scanner models and imaging sites ^12^, making it difficult to incorporate into clinical decision-making.

A reliable quantitative biomarker could facilitate consistent interpretation, particularly in longitudinal MRI-directed follow-up settings such as active surveillance (AS), where distinguishing true biological change from measurement variability is clinically important ^13^. This need is increasingly relevant as AS is widely used for low-risk prostate cancer and is increasingly considered for selected patients with favorable intermediate-risk or GG2 disease, with MRI playing an expanding role in patient selection and follow-up^2,13–15^.

Restriction Spectrum Imaging (RSI) has been introduced as an advanced diffusion MRI technique that enables more refined characterization of tissue microstructure ^16,17^. RSI models the diffusion signal as contributions from four distinct tissue compartments, including restricted intracellular water (RSI-C_1_), hindered extracellular water (RSI-C_2_), freely diffusing water (RSI-C_3_), and vascular flow (RSI-C_4_). A quantitative biomarker for prostate cancer, the RSI restriction score (RSIrs), can be calculated from as little as 2-3 minutes of diffusion data from a standard clinical scanner. Prior studies have shown that RSIrs may outperform conventional diffusion metrics in patient-level cancer detection and may also serve as a valuable adjunct to radiologist interpretation ^18–23^. RSIrs is being studied in an ongoing multi-center, international trial for prostate cancer diagnosis and in a randomized controlled trial for targeted radiotherapy ^24,25^.

In our previous work, we evaluated the reproducibility of prostate RSIrs and ADC measurements within the same scan session, using repeated acquisitions separated by only a few minutes ^26^. While RSIrs demonstrated relatively strong reproducibility, notably higher than that of ADC, we found that diffusion-derived biomarkers may still exhibit non-negligible variability even under highly controlled conditions within the same exam. This observation motivated the present study: to understand the reproducibility of RSIrs and ADC under more clinically realistic conditions, including across different scan sessions, across scanner vendors, and under varying acquisition settings within the same vendor platform. Such analyses are critical for determining the reliability of diffusion-based quantitative biomarkers when used in longitudinal studies or multi-institutional clinical workflows. We also investigate the reproducibility of these biomarkers in patients with prostate cancer characterized by high metastatic potential, referred to as unfavorable-histology prostate cancer (uhPC). Recent studies have identified unfavorable histologic patterns as key clinical risk factors for metastasis and adverse clinical outcomes ^13,27–29^. In this context, uhPC is defined as either high-grade prostate cancer (GG≥3) or intermediate-grade disease that exhibits cribriform morphology, including intraductal carcinoma. Evaluating biomarker reproducibility in this clinically high-risk subgroup is particularly important because reliable imaging biomarkers could play a critical role in risk stratification, treatment planning, and longitudinal disease monitoring.

## Materials and Methods

### Study Population

This prospective study (NCT04992728) was approved by the Institutional Review Board of the University of California San Diego. All participants provided written informed consent. Participants with suspected or known prostate cancer (active surveillance or pre-treatment) were enrolled between August 2022 and January 2026 if they were 18 years or older and able to complete two MRI exams that included an RSI protocol. The first exam includes PI-RADS v2.1 compliant multi-parametric MRI protocol, in addition to RSI, with T2-weighted imaging, DWI and DCE was generally performed as part of the patient’s standard of care either at the University of California San Diego or a community imaging center ^30^. The second exam was performed during a separate appointment on a different scanner at the University of California San Diego, which is dedicated for research. Participants with incomplete exams or poor-quality images were excluded from further analysis. Prostate cancer diagnosis was confirmed on histopathology per clinical routine.

### Data Acquisition and Processing

Acquisition of RSI images required 2-3 minutes per patient. RSI is included in the standard clinical mpMRI protocols at the University of California San Diego and other centers, for use with a commercial, FDA-cleared product. The second exam was exclusively for research and included only RSI and a high-resolution *T*_*2*_-weighted acquisition for anatomical reference. Supplementary Table 1 lists the image acquisition parameters for the RSI and T2-weighted series used for both exams.

RSI data was first processed to correct for background noise and anatomic distortions from eddy currents, gradient nonlinearities, and *B*_*0*_ inhomogeneity ^31–33^. Multiple acquisitions that were acquired at each *b*-value were averaged together. ADC maps were computed from the averaged RSI volume by fitting a standard mono-exponential signal decay model to images with *b*-values less than or equal than 1,000 s/mm^2 17^. A 4-compartment model of the RSI signal was then fit to the data to estimate the signal contribution from each compartment, using a previously described fitting procedure ^17^. The formula for the RSI model used in this study is shown in the Supplementary Materials, with the specific diffusion coefficient parameters listed in Supplementary Table 2. RSIrs was calculated by normalizing the signal contribution of the first compartment, RSI-C_1_, by the median *T*_*2*_-weighted signal in the prostate. The maximum RSIrs value within the prostate (RSIrs-max) was recorded for each patient.

### Prostate and Lesion Contouring

Automatic segmentation of the prostate gland was performed on the *T*_*2*_-weighted MRI scans from both exams using a validated AI model ^34–36^ to obtain preliminary prostate volumes. These prostate contours were then reviewed and corrected by one of two board-certified genitourinary radiologists with expertise in prostate MRI (5 years of experience, >10 years of experience). That same radiologist also identified and graded any intraprostatic lesions according to the Prostate Imaging Reporting & Data System (PI-RADS) v2.1, using mpMRI (*T*_*2*_-weighted, DWI, ADC, DCE) ^8^. The same lesion contours were used for evaluation of quantitative ADC and RSIrs values. Available pathology and radiology reports were cross-referenced, and lesion contours were generated in accordance with the corresponding pathology findings. For participants with multiple MRI-visible lesions, RSIrs-max and mean ADC were derived from the index lesion, as determined by GG and cribriform status when available; otherwise, lesion selection was based on PI-RADS. When multiple lesions were tied on these criteria, the lesion with the larger volume was selected. Manual editing of contours was performed using MIM (MIM Software Inc, Cleveland, OH, USA).

### Statistical Analyses

RSIrs-max and mean ADC within MRI-visible lesions were visualized as scatter plots for all participants, showing the values for scan 1 and scan 2. Scatter plots were also generated for the patient subgroups that (a) had both exams on MRI scanners from the same manufacturer and (b) had exams on MRI scanners from different manufacturers.

Reproducibility was quantified using the Pearson correlation coefficients, with 10,000 bootstrap resamples used to estimate 95% confidence intervals for the correlations.

For ADC, the within-lesion mean is used in current clinical practice and is therefore our choice for the primary analyses. However, for comparison, we secondarily repeated the main analyses using the minimum and 10^th^ percentile of ADC within lesions.

We note that repeatability coefficient and reproducibility coefficient are two metrics that have been proposed for evaluation of test-retest datasets of imaging biomarkers ^10^. These rely on within-subject standard deviation (wSD) or within-subject coefficient of variation (wCV), approaches well suited for more data with more than two repeated measurements per subject. As our study included only two scans per patient, scatter plots and correlation-based analyses provide a practical approach for evaluating agreement between the paired measurements. A strong correlation between repeated measurements indicates that the biomarker preserves consistent relative ordering across scans, which is an important aspect of reproducibility for quantitative imaging biomarkers, particularly in studies with limited repeated acquisitions.

### Sample Size Determination

Published modeling analyses determined that at least 35 participants are needed to estimate precision of a quantitative imaging biomarker, a result endorsed by the Quantitative Imaging Biomarkers Alliance (QIBA) ^10,37^. We sought to enroll at least 70 participants to permit subgroup analyses.

### Evaluating the impact of zero-filled interpolation on cross-session reproducibility

Over the course of the study, we noted that RSIrs values can vary between images that are reconstructed with or without zero-filled interpolation (a setting that some vendors use by default and others do not), due to differences in the amount of partial volume artifacts in the prostate. To assess how zero-filled interpolation affects the reproducibility of mean ADC and RSIrs-max within MRI-visible lesions, we compared the correlation between measurements obtained using zero-filling for both scans to those obtained using zero filling for only one scan.

## Results

### Study Population

Figure 1 outlines the inclusion and exclusion of participants enrolled in this study. Overall, 61 participants met all the criteria for inclusion. Table 1 provides a comprehensive summary of demographic and diagnostic information on the included participants. Exams were conducted using three scanner models from GE Healthcare (Discovery MR750, Discovery MR750w, and Signa Premier) and one from Siemens Healthineers (Prisma). A list of scanner models by imaging center is provided in Supplementary Table 3. Of the 61 included participants, 58 had an MRI-visible lesion that could be contoured. Of these 58, 20 were scanned on machines from the same manufacturer for both exams and 38 were scanned on machines from different manufacturers. uhPC was found in 26 of the 58 participants with visible lesions, of whom 10 were scanned on machines from the same manufacturer for both exams and 16 were scanned on machines from different manufacturers. The protocol for 20 participants included two sets of RSI scans, one reconstructed with zero-filled interpolation and one without.

**Table 1.** Demographic and diagnostic summary of the participant cohort included in this study.

| Characteristic |  |
| --- | --- |
| No. of cases | 61 |
| No. of cases with uhPC | 26 |
| Time from first MRI to second MRI (days)* | 32 (11-65) |
| Age (years)* | 69 (63-74) |
| PSA (ng/ml)* | 6.8 (4.8-9.7) |
| Prostate Volume (ml)* | 50.9 (38.2-65.2) |
| PSA Density (ng/ml<sup>2</sup>)* | 0.12 (0.08-0.24) |
| Scanner vendor between exams |  |
| Same Vendor | 22 |
| Different Vendor | 39 |
| PI-RADS score |  |
| 1 | 0 |
| 2 | 0 |
| 3 | 4 |
| 4 | 24 |
| 5 | 30 |
| No Score | 3 |
| Gleason Grade Group |  |
| Benign | 3 |
| 1 | 11 |
| 2 | 23 |
| 3 | 9 |
| 4 | 3 |
| 5 | 9 |
| No Biopsy | 3 |
| Biopsy Pathology |  |
| Systematic biopsy only | 12 |
| Systematic and targeted biopsy | 46 |
| No biopsy within 6 months of MRI scan | 3 |
| Self-Reported Ethnicity |  |
| Hispanic | 6 |
| Non-Hispanic | 54 |
| Unknown | 1 |
| Self-Reported Race |  |
| Asian | 8 |
| Black or African American | 4 |
| White | 42 |
| Other / Unknown | 7 |

\* Data are median with IQR in parentheses.
uhPC = unfavorable-histology Prostate Cancer (Gleason Grade Group = 2 with cribriform or intraductal feature and Gleason Grade Group >= 3).

**Figure 1.**
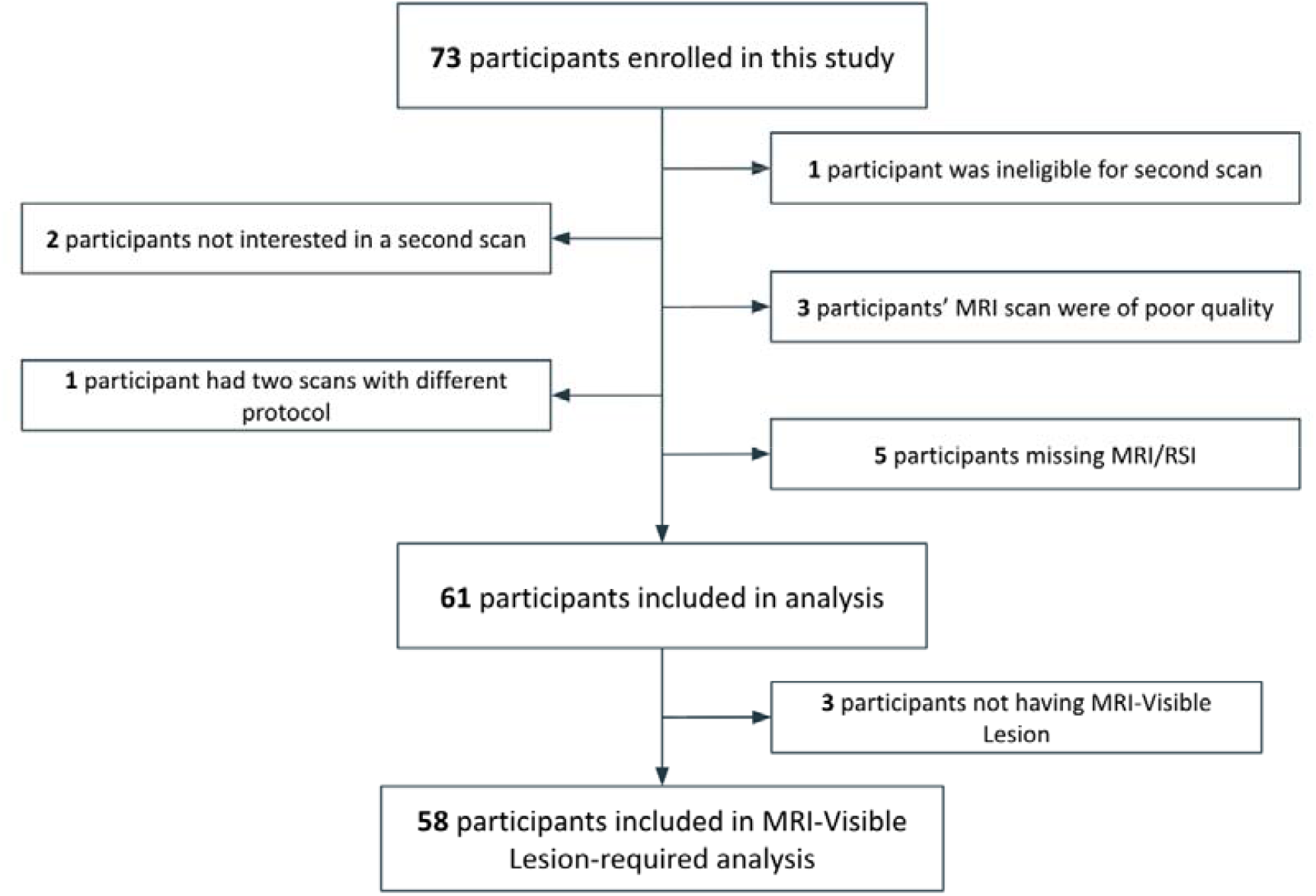
Flowchart showing the inclusion and exclusion criteria of participants enrolled in this study.

### Evaluating cross-session reproducibility within MRI-visible lesions

Figure 2(A) shows scatter plots evaluating cross-session reproducibility of mean ADC within MRI-visible lesions. For participants with MRI-visible lesions (n=58), the correlation for mean ADC was 0.55 (95% CI: 0.23-0.76). For participants scanned on the same scanner vendor across both sessions (n=20), the correlation was 0.76 (95% CI: 0.27-0.95), whereas for participants scanned on different scanner vendors across sessions (n=38), the correlation was 0.31 (95% CI: −0.07-0.62).

**Figure 2.**
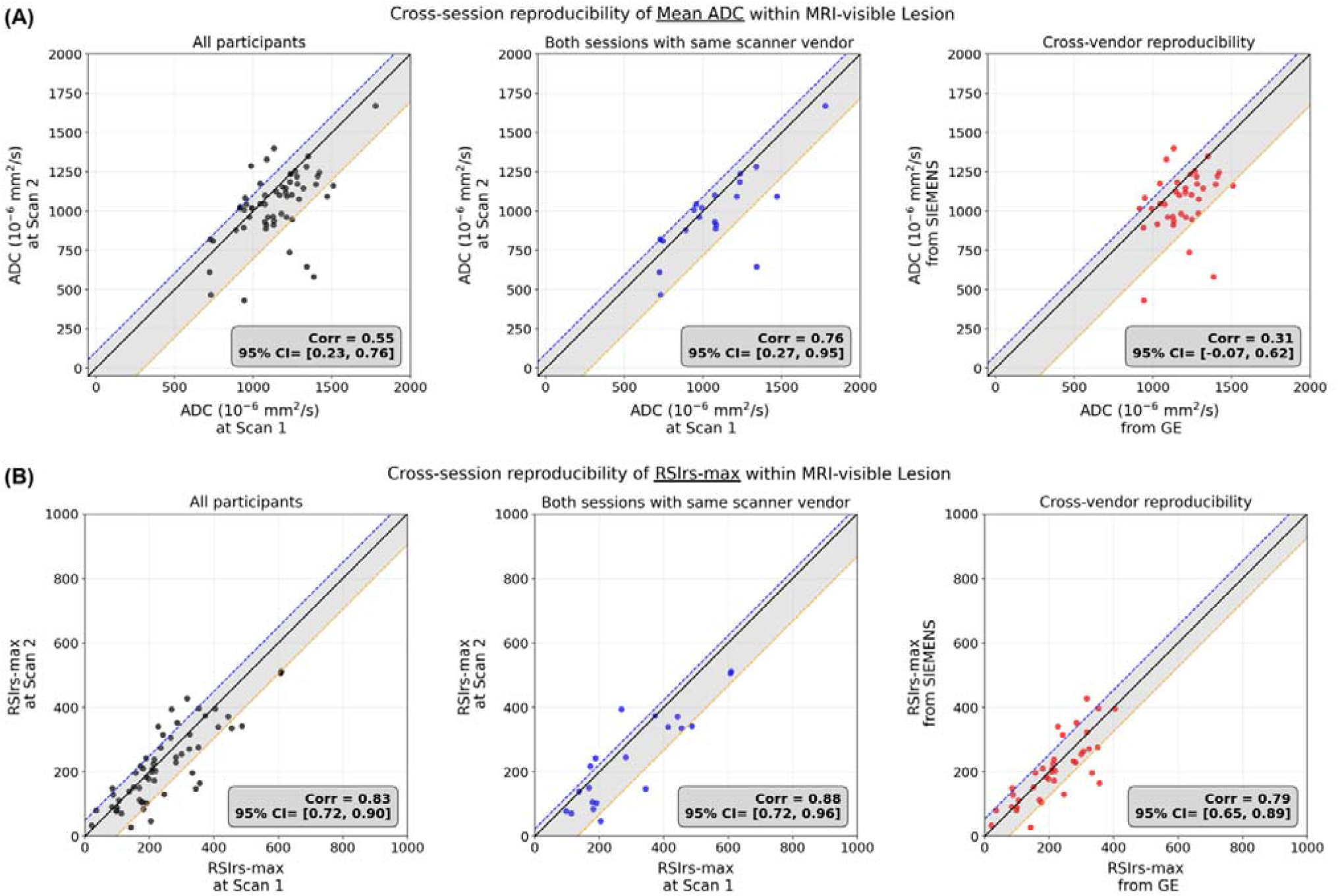
Scatter plots for cross-session reproducibility of RSIrs-max and Mean ADC within MRI-Visible Lesion. The shaded region represents the mean ± 1 standard deviation (SD) of the difference (y − x), and the dashed lines indicate the mean difference ± 1 SD. Corr denotes the correlation between x and y, and the corresponding 95% bootstrap confidence interval (95% CI), based on 10,000 resamples, is also provided. **(A)** Cross-session reproducibility of Mean ADC within MRI-Visible Lesion. **(B)** Cross-session reproducibility of RSIrs-max within MRI-Visible Lesion.

Figure 2(B) shows scatter plots evaluating cross-session reproducibility of RSIrs-max within MRI-visible lesions. For all participants with MRI-visible lesions (n=58), the correlation for RSIrs-max was 0.83 (95% CI: 0.72–0.90). For participants scanned on the same vendor across both sessions (n=20), the correlation was 0.88 (95% CI: 0.72–0.96), and for participants scanned on different vendors across sessions (n=38), the correlation was 0.79 (95% CI: 0.65–0.89).

ADC reproducibility was not substantially improved when using the minimum or 10^th^ percentile instead of the mean (Supplementary Figure 1).

### Evaluating cross-session reproducibility within MRI-visible lesions for participants with uhPC

Figure 3(A) shows scatter plots evaluating cross-session reproducibility of mean ADC within MRI-visible lesions among participants with uhPC. For all uhPC participants with MRI-visible lesions (n=26), the correlation for mean ADC was 0.42 (95% CI: −0.02–0.83). For participants scanned on the same vendor across both sessions (n=11), the correlation was 0.29 (95% CI: −0.51–0.99), whereas for participants scanned on different vendors across sessions (n=15), the correlation was 0.44 (95% CI: 0.08–0.74).

**Figure 3.**
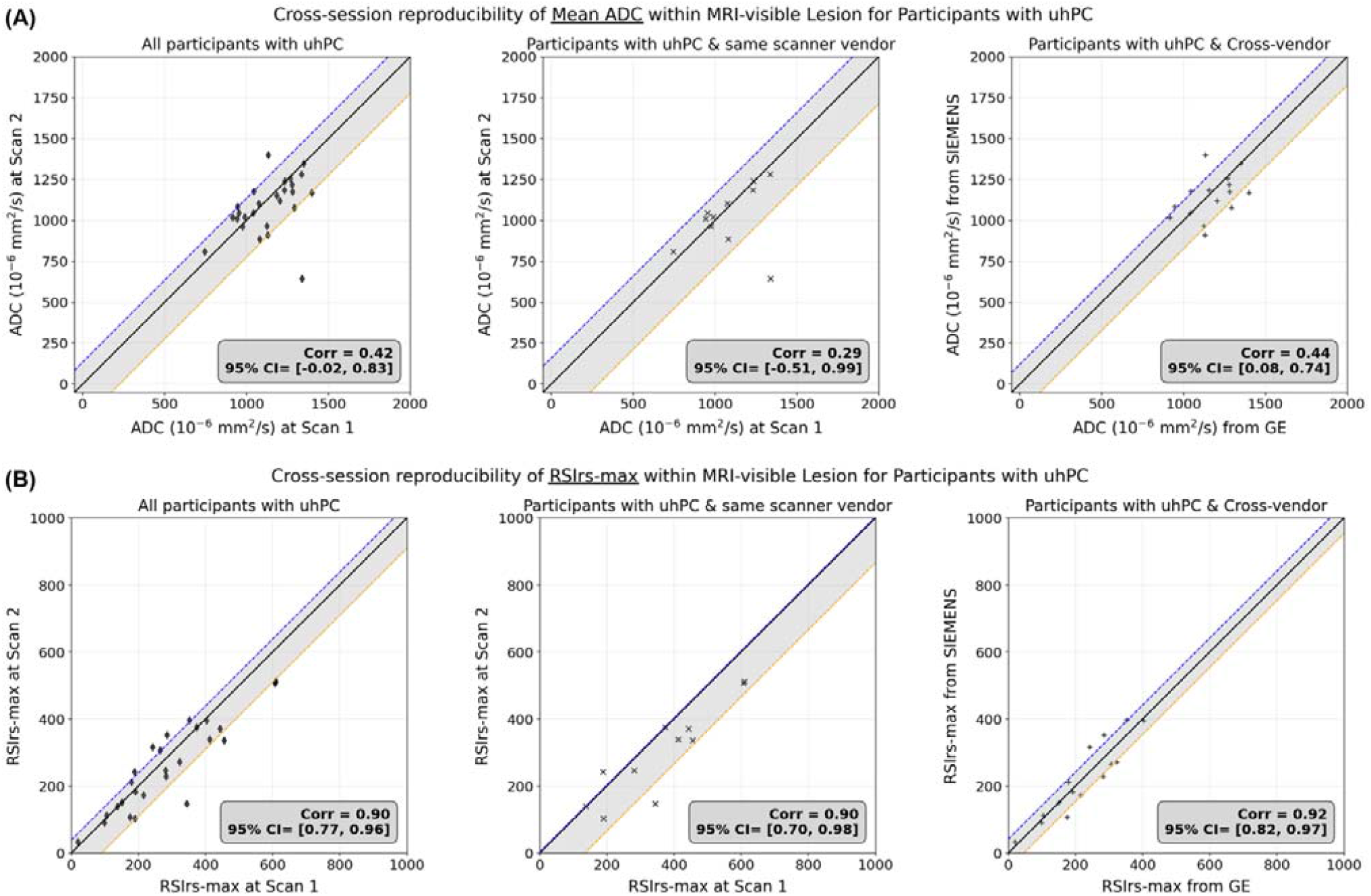
Scatter plots for cross-session reproducibility of RSIrs-max and Mean ADC within MRI-Visible Lesion for Participants with uhPC. The shaded region represents the mean ± 1 standard deviation (SD) of the difference (y − x), and the dashed lines indicate the mean difference ± 1 SD. Corr denotes the correlation between x and y, and the corresponding 95% bootstrap confidence interval (95% CI), based on 10,000 resamples, is also provided. **(A)** Cross-session reproducibility of Mean ADC within MRI-Visible Lesion for Participants with uhPC. **(B)** Cross-session reproducibility of RSIrs-max within MRI-Visible Lesion for Participants with uhPC.

Figure 3(B) shows scatter plots evaluating cross-session reproducibility of RSIrs-max within MRI-visible lesions among participants with uhPC. For all uhPC participants with MRI-visible lesions (n=26), the correlation for RSIrs-max was 0.90 (95% CI: 0.77–0.96). For participants scanned on the same vendor across both sessions (n=11), the correlation was 0.90 (95% CI: 0.70–0.98). For participants scanned on different vendors across sessions, the correlation was 0.92 (95% CI: 0.82–0.97).

### Evaluating the impact of zero-filled interpolation on cross-session reproducibility

Figure 4 (A) shows scatter plots evaluating reproducibility of mean ADC within MRI-visible lesions, comparing measurements with and without zero-filled interpolation acquired within the same scanning session. The correlation between exams with and without interpolation was 0.32 (95% CI: −0.24 to 0.72). The correlation between two exams with interpolation was 0.72 (95% CI: 0.35 to 0.88). The correlation between two scans acquired during a single exam with and without interpolation was 0.69 (95% CI: 0.37 to 0.88).

**Figure 4.**
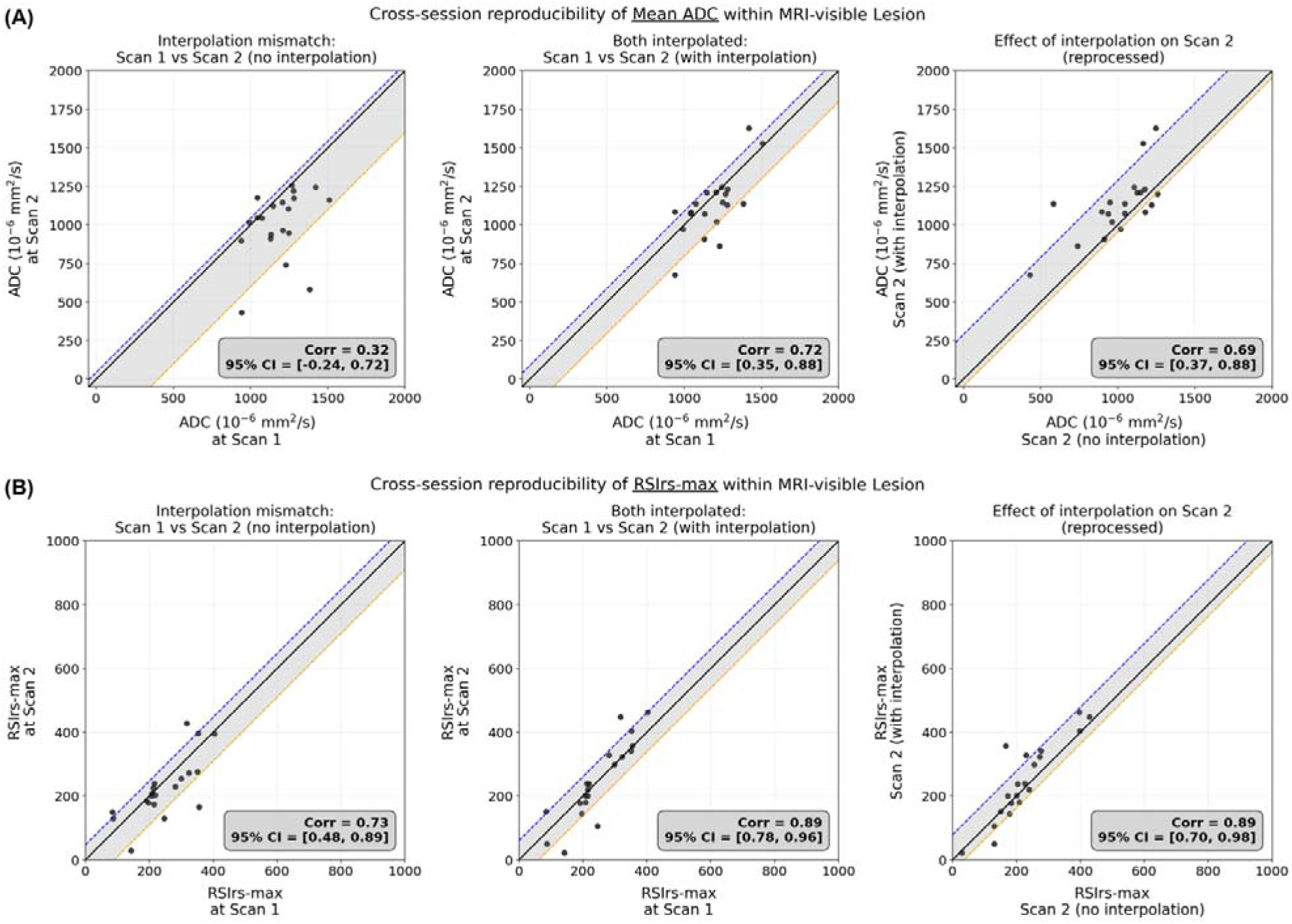
Scatter plots for cross-session reproducibility of RSIrs-max and Mean ADC within MRI-Visible Lesion across vendors, with and without interpolation. The shaded region represents the mean ± 1 standard deviation (SD) of the difference (y − x), and the dashed lines indicate the mean difference ± 1 SD. Corr denotes the correlation between x and y, and the corresponding 95% bootstrap confidence interval (95% CI), based on 10,000 resamples, is also provided. (**A)** Cross-session reproducibility of Mean ADC within MRI-Visible Lesion across vendors, with and without interpolation. **(B)** Cross-session reproducibility of RSIrs-max within MRI-Visible Lesion across vendors, with and without interpolation.

Figure 4 (B) presents the corresponding analysis for RSIrs-max. The correlation between exams with and without interpolation was 0.73 (95% CI: 0.48 to 0.89), increasing to 0.89 (95% CI: 0.78 to 0.96) between two exams with interpolation. The correlation between two scans acquired during a single exam with and without interpolation was 0.89 (95% CI: 0.70 to 0.98).

## Discussion

While PI-RADS provides a standardized approach for image interpretation, it is based on qualitative assessments that can differ even among expert subspecialist radiologists ^30^. In this study, we evaluated the reproducibility of two diffusion-based biomarkers (mean ADC ^38^ and RSIrs-max) under clinically realistic conditions. We found that mean ADC showed lower reproducibility, which may limit its use as a quantitative biomarker. This is why mean ADC is typically used only semi-quantitatively as part of an overall subjective interpretation according to PI-RADS v2.1 ^8^. Across all our analyses, RSIrs-max demonstrated consistently better reproducibility than mean ADC. This pattern was observed both when scans were acquired on the same scanner and when they were acquired on scanners from different manufacturers. Notably, a maximum value is inherently more sensitive to noise and measurement variability than a mean, but RSIrs-max nonetheless maintained superior reproducibility compared to mean ADC ^39^.

We also evaluated reproducibility within the MRI-visible lesions of participants with uhPC, a subgroup associated with higher metastatic potential for whom accurate imaging biomarkers could provide the greatest clinical benefit. Recent work has highlighted the potential prognostic value of ADC in prostate cancer ^11^. However, we found mean ADC showed limited reproducibility, suggesting that it may be less suitable as a standalone quantitative metric for longitudinal assessment in this setting. RSIrs-max, on the other hand, had correlation coefficients ≥0.90 for within-vendor and between-vendor comparisons in participants with uhPC. Such stability in a quantitative biomarker is particularly important for applications involving longitudinal monitoring, treatment response assessment, and multi-center clinical studies ^40^.

Finally, we observed that consistent application of zero-filled interpolation improves the reproducibility of RSIrs-max. Agreement between RSIrs-max measurements between exams was higher when interpolation was applied consistently for both RSI scans instead of only one. This finding likely resulted from partial volume artifacts on the images without interpolation, which reduce the RSI signal in lesions due to signal averaging with adjacent non-cancerous tissue. By mitigating the partial volume averaging between voxels ^41^, zero-filled interpolation enables better isolation of the diffusion signal from lesions and more reliable estimation of RSIrs values. These results highlight the importance of preprocessing and standardization techniques in improving cross-vendor reproducibility of diffusion-based imaging biomarkers.

In a previous study ^26^, we evaluated *intra-session repeatability* using two scans acquired within the same imaging session. The correlation of RSIrs-max within the prostate for participants who underwent two scans during the same session (Supplementary Figure 1) was comparable to the correlation observed in the present study for participants in the present study who underwent two scans across different sessions using scanners from the same manufacturer (Supplementary Figure 2). In other words, the reproducibility of RSIrs observed between scans performed in separate sessions approximately matches the repeatability observed between two acquisitions in the same session within approximately a 10-minute interval.

There are several technical factors that can contribute to imperfect repeatability with MRI scans, including even slightly different positioning of the patient with respect to the magnetic field and receiver coils, temperature-related drift in gradient performance, recalibration differences, and shifts in magnetic field heterogeneity. A prior study showed these factors can, together, contribute to modest variation in ADC simply by having the patient get off and back on the scanner ^42^. Factors such as partial-volume effects, slice selection, and index lesion boundary placement may also contribute to variability in measured values between scans. However, because the same index lesion contours were used for both RSI and ADC in the present study, and reproducibility of each metric was directly compared, lesion-related factors do not account for the observed differences between biomarkers.

This study has several limitations. First, the cohort was derived from a limited number of institutions and scanner vendors. Multi-center, multi-vendor validation is needed to assess robustness across diverse clinical settings. Reassuringly, a retrospective multi-center study found RSIrs to be robust to scanner, protocol, and center variation ^40^. Second, each patient underwent only two scans, which precluded estimation of variance-based repeatability and reproducibility metrics ^10^. Finally, this study focused specifically on reproducibility, which is important for longitudinal assessment and quantitative interpretation. Other clinically relevant characteristics, such as diagnostic performance, prognostic value, and treatment-response assessment are of interest and under active investigation^21,24,25,43,44^.

## Conclusion

The commonly used ADC metric, even as a lesion mean value, can vary considerably when the same patient undergoes repeat imaging on different days. RSIrs-max, on the other hand, demonstrated strong reproducibility and consistently outperformed ADC across both same-scanner and cross-vendor settings. These findings remained robust in participants with uhPC and under different vendor interpolation settings. Notably, RSIrs-max reproducibility across different scan sessions on the same scanner is comparable to that observed for two acquisitions obtained only minutes apart within the same session. RSIrs-max is being evaluated in prospective trials for initial diagnosis (NCT06579417; NCT05882253), radiotherapy treatment targeting (NCT06990542), and treatment response assessment (NCT04349501) ^25^.

## Data Availability

All data produced in the present study are available upon reasonable request to the authors

