## Supplementary Materials for "Reproducibility of Apparent Diffusion Coefficient and Restriction Spectrum Imaging Restriction Score in the Prostate Across MRI Sessions, Vendors, and Acquisition Settings: a Prospective Study"

**Supplementary Materials – Info & Tables**

**RSI model equation**

The RSI model equation shows the relationship between the measured DWI signal intensity to the signal contribution from each of four tissue compartments: $S\left( b \right)$ represents the measured DWI signal intensity at a specific *b*-value.

$S\left( b \right) = \sum_{i = 1}^{4} C_{i}e^{-bD_{i}}$	(1)

Where $S\left( b \right)$ represents the measured DWI signal intensity at a specific *b*-value, $C_{i}$ is the signal contribution of a particular compartment to the overall signal, to be determined through model-fitting. $D_{i}$ is the diffusion coefficient that is empirically determined for each of the four compartments. The detailed parameter for each RSI compartments is shown in Supplementary table 1.

| **UCSD Health** | **RSI** | ***T_2_*-weighted** |
| --- | --- | --- |
| Pulse sequence | Diffusion-weighted EPI | Fast Spin Echo (FSE) |
| TR (ms) | 3800 | 3344 |
| TE (ms) | 80 | 102 |
| FOV (mm) | 160 x 160 | 160 x 160 |
| Matrix [resampled dimensions] | 64 x 64 [256 x 256] | 360 x 224 [512 x 512] |
| Slices | 32 | 32 |
| Slice Thickness (mm) | 3 | 3 |
| b-values (s/mm^2^) [number of samples] | 0 [5], 100 [6], 800 [12], 1400 [12], 2500 [18] | N/A |
| Field Strength (T) | 3 | 3 |
| **Outside Imaging Center** | **RSI** | ***T_2_*-weighted** |
| Pulse sequence | Diffusion-weighted EPI | Fast Spin Echo (FSE) |
| TR (ms) | 4800 | 3500 |
| TE (ms) | 87 | 170 |
| FOV (mm) | 180 x 90 | 240 x 240 |
| Matrix [resampled dimensions] | 90 x 46 [256 x 256] | 448 x 320 [512 x 512] |
| Slices | 24 | 32 |
| Slice Thickness (mm) | 4 | 3 |
| b-values (s/mm^2^) [number of samples] | 0 [1], 500 [8], 1000 [8], 2000 [16] | N/A |
| Field Strength (T) | 3 | 3 |

Supplementary Table 1. MRI acquisition parameters for each cohort. All RSI and T2-weighted scans were acquired in the axial plane. UCSD Health = University of California San Diego Health. TR = repetition time. TE = echo time. FOV = field-of-view. FSE = fast spin echo. EPI = echo-planar imaging. RSI = Restriction Spectrum Imaging.

| **RSI compartment** | **Fixed Diffusion Coefficient (s/mm^2^)** |
| --- | --- |
| Restricted Diffusion $\left( C_{1} \right)$ | $1.1 \times{10}^{-4} \left( D_{1} \right)$ |
| Hindered Diffusion $\left( C_{2} \right)$ | $1.8 \times{10}^{-3} \left( D_{2} \right)$ |
| Free Diffusion $\left( C_{3} \right)$ | $3.6 \times{10}^{-3} \left( D_{3} \right)$ |
| Vascular Flow $\left( C_{4} \right)$ | $0.1220 \left( D_{4} \right)$ |

Supplementary Table 2. Description and diffusion coefficient parameter for each of the four RSI model compartments.

| **Institution** | **Scanner models** |
| --- | --- |
| UCSD Health | GE Healthcare Discovery MR750, GE Healthcare Signa Premier, SIEMENS Prisma |
| Outside Imaging Centers | GE Healthcare Discovery MR750w |

Supplementary Table 3. Scanner models used at each imaging center. UCSD Health = University of California San Diego Health.

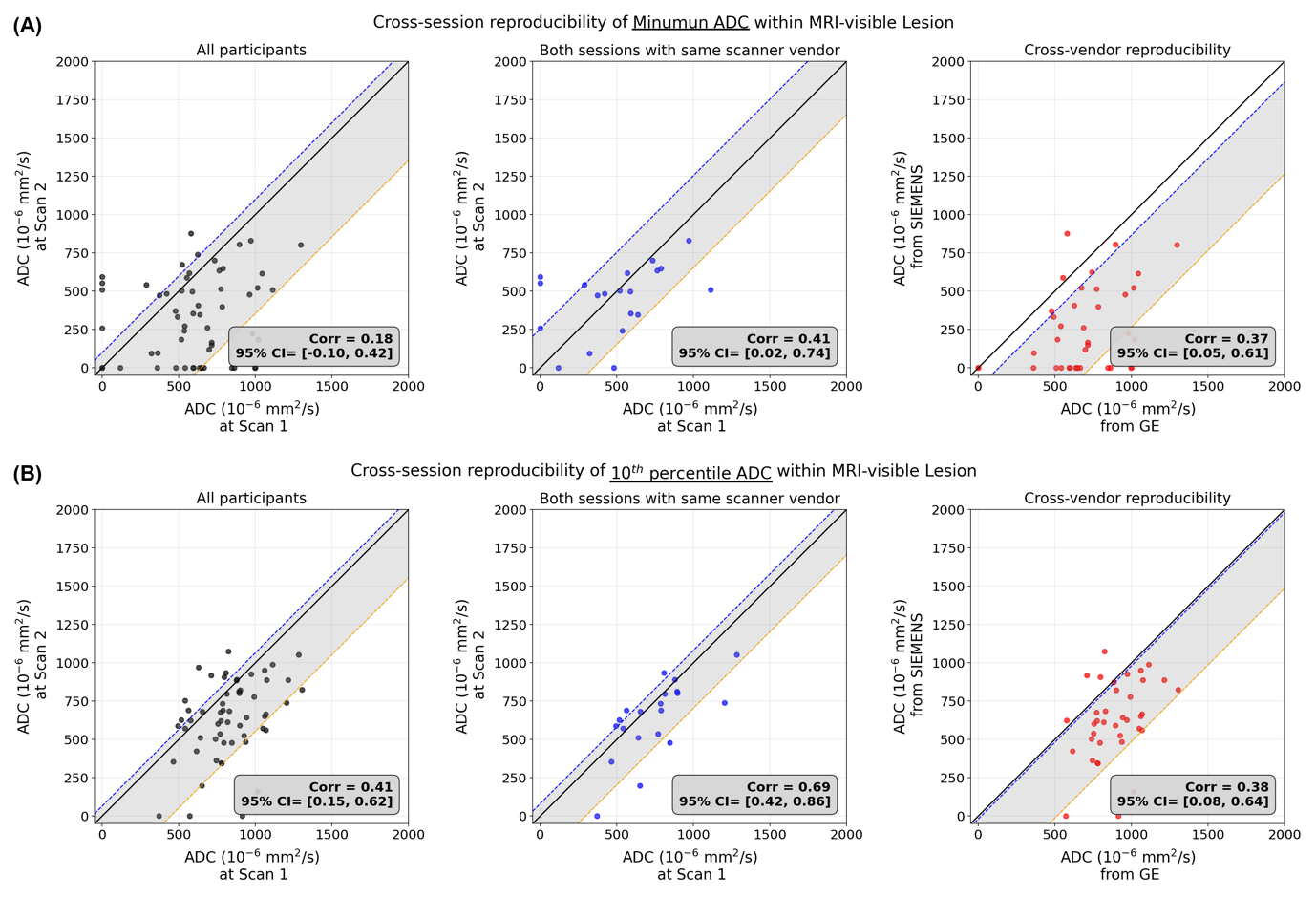

Supplementary Figure 1. Scatter plots for cross-session reproducibility of Minimum and 10^th^ percentile ADC within MRI-Visible Lesion. The shaded region represents the mean ± 1 standard deviation (SD) of the difference (y − x), and the dashed lines indicate the mean difference ± 1 SD. Corr denotes the correlation between x and y, and the corresponding 95% bootstrap confidence interval (95% CI), based on 10,000 resamples, is also provided. **(A)** Cross-session reproducibility of Minimum ADC within MRI-Visible Lesion. **(B)** Cross-session reproducibility of 10^th^ percentile ADC within MRI-Visible Lesion.

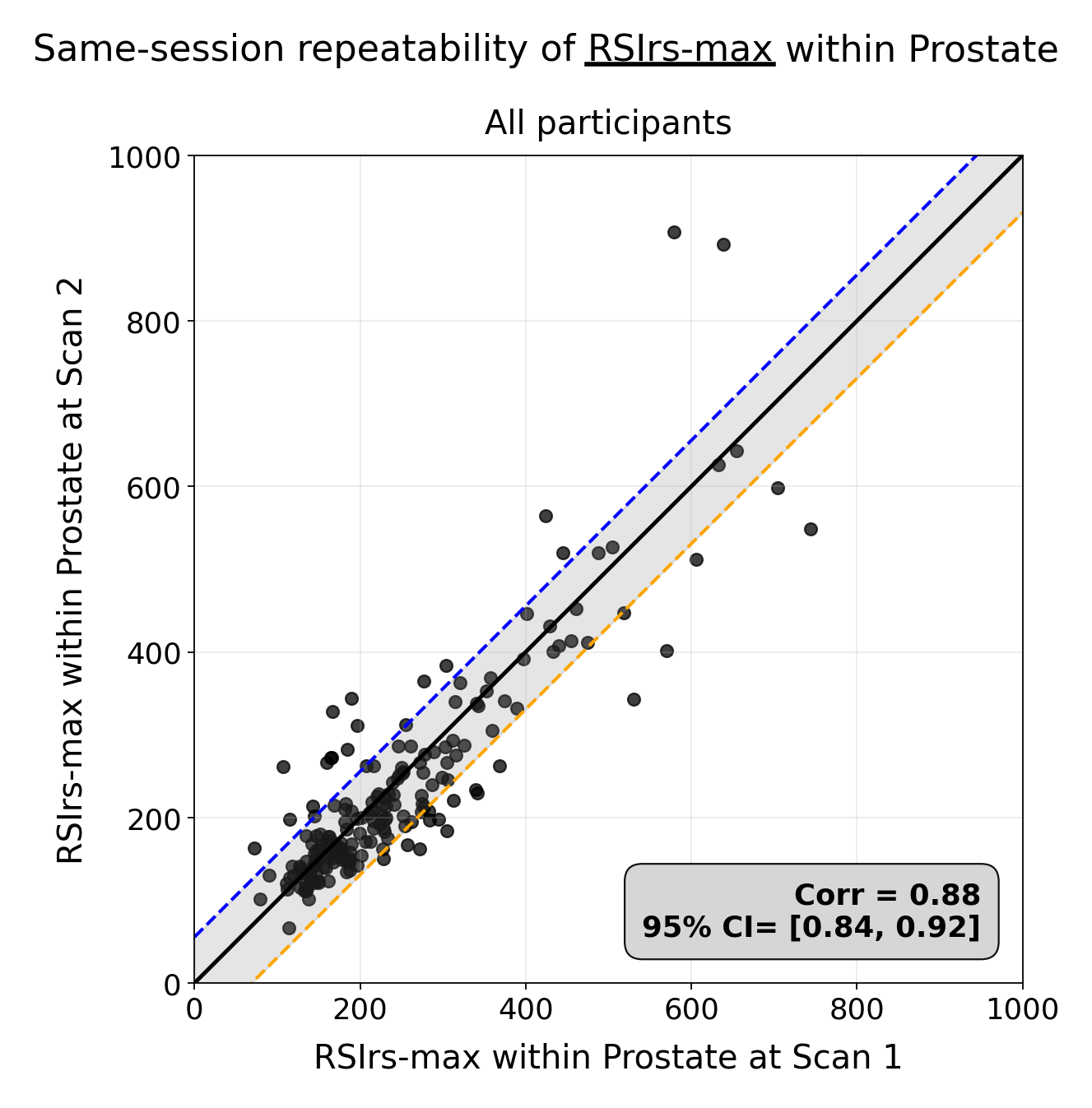

Supplementary Figure 2. Scatter plot for same-session reproducibility of RSIrs-max within the entire prostate (lesion and non-lesion). This approach has been successfully used in previous studies to evaluate the probability of clinically significant or aggressive prostate cancer without the need of an expert-defined lesion. The shaded region represents the mean ± 1 standard deviation (SD) of the difference (y − x), and the dashed lines indicate the mean difference ± 1 SD. Corr denotes the correlation between x and y, and the corresponding 95% bootstrap confidence interval (95% CI), based on 10,000 resamples, is also provided.

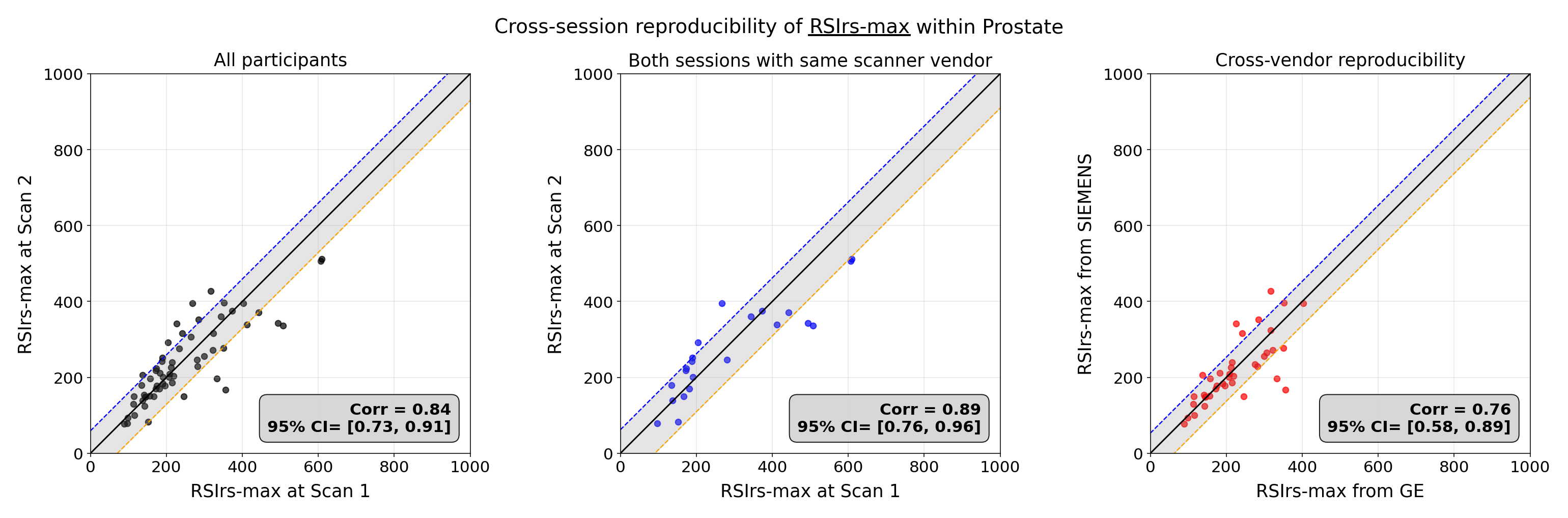

Supplementary Figure 3. Scatter plots for cross-session reproducibility of RSIrs-max within prostate. The shaded region represents the mean ± 1 standard deviation (SD) of the difference (y − x), and the dashed lines indicate the mean difference ± 1 SD. Corr denotes the correlation between x and y, and the corresponding 95% bootstrap confidence interval (95% CI), based on 10,000 resamples, is also provided.
